# Systematic Review Protocol: The Effects of Theory-Based Interventions for Self-Help Practices in the Management of Mild to Moderate Depression

**DOI:** 10.1101/2024.12.20.24319466

**Authors:** Asma’ Khalil, Zahir Izuan Azhar, Norley Shuib, Philip Baker, Chen Xin Wee

## Abstract

**Background:** Depression is a leading global cause of disability, significantly affecting individuals’ quality of life and contributing to a substantial worldwide health burden. Self-help practices have emerged as cost-effective and scalable strategies for managing mild to moderate depression, empowering individuals to take an active role in their mental health care. Theory-based interventions, which are grounded in established psychological frameworks, provide a systematic approach to designing, implementing and evaluating these self-help practices. However, despite the growing availability of self-help interventions, their effectiveness varies, and many lack a solid theoretical foundation. Summarized evidence of the effects of theory-based interventions remains limited.

**Objectives:** This systematic review aims to evaluate the effects of theory-based interventions incorporating self-help practices in reducing depressive symptoms severity among adults with mild to moderate depression.

**Methods:** The review will follow PRISMA guidelines and has been registered with the International Prospective Register of Systematic Reviews (PROSPERO). Randomised trials evaluating theory-based interventions, such as Cognitive Behavioral Therapy (CBT), Acceptance and Commitment Therapy (ACT), and Mindfulness-Based Interventions (MBIs), delivered via digital platforms will be included. Studies will be identified through searches of PubMed, Scopus, Web of Science, Science Direct and Cochrane bibliographic databases. Data extraction and risk of bias assessment will be conducted independently by two reviewers, focusing on study characteristics, intervention details and critical outcomes (reduction in depressive symptom severity and adverse effects).

**Results:** The review will synthesize evidence on the effects and adaptability of theory-based interventions, highlighting the specific contributions of theoretical frameworks to intervention outcomes. Findings will be presented in tabular and narrative formats, identifying gaps and implications for clinical practice and future research.

**Conclusion:** This systematic review will provide actionable insights into the design and implementation of theory-based interventions for managing mild to moderate depression. The findings aim to inform evidence-based guidelines for primary care integration, promoting accessible, scalable and effective mental health solutions globally.

## 1. Introduction

Depression remains one of the leading global health concerns, contributing significantly to the overall disease burden. According to the World Health Organization (WHO), it affects over 280 million people globally, making it a top contributor to global disability in 2022 (1). Primary care settings play a crucial role as the first point of contact for individuals experiencing depressive symptoms, thereby emphasizing the importance of effective and accessible treatment options within this context (2). While commonly employed, pharmacological treatments come with various challenges, including adherence issues and potential side effects. In response to these challenges, theory-based interventions have emerged as a non-pharmacological approach to managing depression (3).

Theory-based interventions in mental health are structured approaches rooted in psychological, behavioral, or social theories. These interventions target specific psychological mechanisms to influence attitudes, beliefs and behaviors, promoting sustainable change in individuals (4). By integrating evidence-based practices, such interventions improve mental health treatment’s predictability, adaptability and effectiveness (5). Self-help practices in mental health encompass strategies individuals employ independently to enhance their emotional and psychological well-being. Theory-based self-help interventions play a crucial role in treating depression, as they provide a structured framework for designing, implementing and evaluating interventions effectively (6).

Several theories are commonly used in advising self-help practices. For example, Cognitive Behavioral Therapy (CBT) is embedded in cognitive theory and focuses on identifying and restructuring maladaptive thought patterns and behaviors. CBT has demonstrated efficacy across various conditions, including anxiety, depression and post-traumatic stress disorder (PTSD) (7). Similarly, motivational interviewing, informed by self-determination theory, aims to enhance intrinsic motivation for behavior change, particularly in substance abuse and chronic health conditions (8). Other examples include Mindfulness-Based Interventions (MBI) focus on fostering conscious, non-judgmental awareness of the present moment to enhance health and well-being (9), and family systems therapy, emphasizes understanding and addressing complex relational dynamics within families, which influence individual behaviors and relationships (10).

The emergence of digital health innovations, including e-health and mHealth platforms, has revolutionized the mental health care delivery. E-health refers to online platforms for healthcare services, while mHealth leverages mobile technology to provide interventions (11). These platforms offer privacy, flexibility and scalability, addressing barriers such as stigma and limited access to care, which are common challenges for individuals with depression (12). Evidence supports the role of digital interventions in improving access, adherence and outcomes for managing mild to moderate depression (13).

## 2. Rationale

Depression, particularly in its mild form, can significantly impact individuals’ quality of life. Theory-based non-pharmacological interventions, grounded in established psychological frameworks, could serve as effective modalities for managing mild to moderate depressive symptoms. Such interventions could provide valuable support to individuals awaiting specialist referral or as an alternative to pharmacological treatments, promoting accessibility and patient autonomy (14).

Currently, there is an increasing availability of self-help interventions, particularly through digital platforms, aimed at mitigating depressive symptoms. While these interventions promise scalability and cost-effectiveness, emerging evidence suggests that their efficacy may vary widely, with many failing to demonstrate consistent or significant benefits in reducing depressive symptoms (15). Given this, it is essential to comprehensively evaluate the impact of self-help practices on reducing depressive symptoms. This includes exploring the methods, theoretical underpinnings, and intensity of interventions to understand their true effectiveness. Despite the potential benefits of theory-based approaches, evidence supporting their implementation and efficacy in self-help interventions remains scarce (16).

To address this gap in knowledge, we propose a systematic review that aims to evaluate the effects of theory-based interventions for self-help practices in the management of mild to moderate depression. By synthesizing recent evidence from randomised trials, this review seeks to provide a comprehensive understanding of how theory-based interventions for self-help practices compare to pharmacological treatments and other therapeutic approaches in alleviating depressive symptoms and preventing relapse. The findings will inform the development of evidence-based guidelines and provide insights into optimizing the integration of digital health tools into primary care and community-based mental health services.

## 3. Objectives

This systematic review aims to achieve the following objectives:

i. To evaluate the effects of theory-based interventions for self-help practices aimed at reducing depressive symptoms severity among adults with mild to moderate depression.
ii. To assess the theory-based interventions for self-help practices to be adopted in primary healthcare settings to manage of adults with mild to moderate depression.

## 4. Methods

### 4.1. Design and registration

This protocol has been registered with the International Prospective Register of Systematic Reviews (PROSPERO) under registration number CRD42024613188. This systematic review will follow the Preferred Reporting Items for Systematic Reviews and Meta-Analyses (PRISMA) guidelines (17). The review will focus on randomized trials, also known as randomized controlled trials (RCTs) that examine the effects of theory-based interventions for self-help practices in reducing depressive symptoms among adults with mild to moderate depression. The methodological approach includes a structured search strategy, defined eligibility criteria, data extraction and risk of bias assessment procedures.

### 4.2. Information Sources

A comprehensive search will be conducted across bibliographic databases, including PubMed MEDLINE, Scopus, Web of Science, ScienceDirect and Cochrane. The search will include peer-reviewed articles published since 2014 to February 2025 representing contemporary approaches.

### 4.3. Search Strategy

The search will be constructed using keywords and limiters appropriate to the database:

- depress* AND
- (theory OR theoret* OR model* OR principle* OR construct* OR framework*) AND
- (online OR web* OR digital OR mobile OR mHeatlh OR m-health OR e-health OR educat OR intervention OR behavioural OR behavioral) AND
- (self-care OR self-help OR self-manage*)

### 4.4. Eligibility Criteria

The study selection process will be guided by the PICO framework:

i. **People (P):**

- Adults aged 18 years old and more, screened or diagnosed with mild to moderate depression and/ or with depression symptoms.
- Specifically, a formal diagnosis of the major depressive disorder based on the Diagnostic and Statistical Manual of Mental Disorders (DSM-5) or International Classification of Diseases, 10th Revision (ICD-10), or
- Participants screened with validated commonly used screening tools in the field of psychiatry such as Patient Health Questionnaire (PHQ-9), Depression, Anxiety and Stress Scale (DASS-21), Center for Epidemiologic Studies Depression Scale (CES-D), Beck Depression Inventory (BDI) or Hamilton Depression Rating Scale (HDRS).
- The participants can either diagnosed with depression only or with other medical comorbidity that are commonly associated with depression e.g. chronic diseases.
ii. **Intervention (I):** To be eligible, interventions must meet the following criteria:

- The intervention must be theory-based, which may include, but is not limited to, Mindfulness-Based Interventions (MBIs), Cognitive Behavioral Therapies (CBTs), Acceptance and Commitment Therapy (ACT), or Behavioral Activation Therapy (BAT) as the primary intervention.
- The intervention must be delivered either as a standalone approach or as part of an integrated hybrid model, and it must exclude pharmacological treatment.
- The intervention must be administered through non-traditional formats, such as text, audio, video, CD/DVD, smartphone, computer, Internet (e.g., automated emails, web-based applications, automated phone calls, short text messages), or individual exercises like ‘therapeutic writing’.
- The intervention must be designed to be delivered independently of a therapist, providing only limited support, guidance, or assistance.
- The intervention must focus solely on self-help practices for individuals.
- The intervention can be held in clinical or non-clinical settings.
iii. **Comparison (C):**

- Pharmacological treatment, symptoms monitoring, psychosocial intervention, therapist-led psychotherapy and combination treatment.
iv. **Outcomes (O):** Severity of depressive symptoms measured by adverse events. Studies with any of these designs or criteria will be excluded:

- RCTs without ethical approval from an accredited, recognized ethics committee or institutional review board.
- Non-randomised studies, observational studies, case studies or commentaries.
- Studies not measuring depression as an outcome.
- Studies not measuring depression pre-and post-intervention.
- Long-term open, uncontrolled extension follow-up of an RCT.
- Interventions of offering on support or guidance provided in a group format.
v. **Timing:** Outcomes will be measured across two time points to capture the effects of theory-based interventions for self-help practices in managing mild to moderate depression:

- Short-term outcomes: Measured within 0-3 months post-intervention.
- Medium-term outcomes: Measured between 3-6 months post-intervention.

This analysis will provide valuable insights into the durability and lasting impact of the intervention effects.

### 4.5. Study Selection

The search results will be imported into EndNote and duplicate entries will be removed and imported in Rayyan software for shared screening. The study selection process will involve a two-stage screening approach. In the first stage, two independent reviewers (AK, CXW) will screen the titles and abstracts with the assistance of Rayyan’s machine learning capabilities. In the second stage, full texts of potentially eligible studies will be reviewed against the predefined inclusion criteria. Any disagreements between the reviewers are resolved by a third reviewer (ZIA). Reasons for exclusion at this stage will be noted.

A PRISMA flowchart will outline the study selection process, including the number of records identified, screened, excluded and included in the review.

## 5. Data Extraction and Management

Data extraction will be performed using a standardized data extraction form, which will be created in Microsoft Excel. Publications describing the same study will be considered as one study. Two independent reviewers will extract relevant data from each included study. Any disagreements between the reviewers will be resolved through discussion, with the involvement of a third reviewer if necessary. The data to be extracted include:

- Study characteristics: author, publication year, country of conduct, study design.
- Participant characteristics: sample size, age, gender, equity markers and depression severity.
- Diagnostic criteria for depression (DSM-5 or ICD-10) and screening tool used.
- Intervention details: type and intensity of the interventions (18), duration, delivery method (in-person, online), resource usage, and process evaluations.
- Comparison group details: pharmacological treatment, symptoms monitoring, psychosocial intervention, therapist-led psychotherapy and combination treatment.
- Outcome measures and measurement tools: changes in depressive symptoms (e.g., PHQ-9, BD-II or CES-D scale).

### 5.1. Study Characteristics

A summary table will present the key characteristics of the included studies, including:

- Study design (e.g., RCT/ randomised trial/ single-arm intervention).
- Study settings: location, year of study.
- Sample size and participant demographics (place of residence, ethnicity, occupation, gender, religion, education, social capital and socio-economic position).
- Study population/ participants (e.g., adults with chronic diseases, HIV, cancer).
- Diagnostic criteria for depression (DSM-IV/ ICD-10) and screening tool used.
- Details of the intervention: theory-based, duration, delivery method, platform used and resource usage.
- Comparator group(s) (e.g., pharmacological treatment, symptoms monitoring).
- Outcome measurement tool (e.g., PHQ-9, BD-II, DASS-21, HAS, CES-D scale).

### 5.2. Outcomes: Effects of Theory-Based Interventions

#### 5.2.1 Critical Outcomes

The primary outcomes of this systematic review will be the degree to which theory-based interventions effect the severity of depressive symptoms among adults with mild to moderate depression and any adverse effects. The change in depressive symptoms severity will be evaluated through validated assessment tools commonly applied in mental health research. Examples of these tools include:

- PHQ-9: A self-reported measure assessing the severity of depressive symptoms over the past two weeks.
- BDI-II: A widely used 21-item scale for evaluating depressive symptomatology.
- CES-D: A screening instrument that measures depressive symptoms in general populations.
- HDRS: A clinician-administered assessment evaluating symptom severity.

Changes in depressive symptoms will be presented both in terms of:

i. Precision and Statistical Significance: Measured as the mean difference or percentage change in symptom scores from baseline to post-intervention.
ii. Clinical Significance: Defined as a pre-determined threshold of symptom reduction, such as a decrease of at least 50% on a scale or remission (e.g., achieving a PHQ-9 score <5) (19).

Adverse events reported in the trial will be identified and analyzed. This will involve examining any reported adverse effects or unintended consequences, such as worsening symptoms, participant distress, or high dropout rates due to negative experiences. Ensuring the safety of interventions is essential to mitigate risks and build confidence in their use among both healthcare providers and users (14).

#### 5.2.2 Important Outcome

To provide a comprehensive evaluation of self-help interventions for managing depressive symptoms, this systematic review will include health equity consideration as an important outcome. This includes assessing the scalability and adaptability of the interventions across diverse settings, populations and delivery platforms. Factors like ease of integration into existing healthcare systems and the intervention’s flexibility to accommodate cultural, socio-economic and demographic variations will be considered. These aspects are critical to understanding the potential for widespread use and the practicality of implementation in real-world scenarios (15).

### 5.3. Risk of Bias Assessment

The risk of bias of the included studies will be evaluated using the Cochrane Risk of Bias Tool (RoB 2) (20). It is a comprehensive and standardized framework developed by the Cochrane Collaboration to assess the risk of bias in the results of RCTs particularly in the design, conduct and reporting of trials. The risk of bias assessment will be independently conducted by two reviewers (AK, CXW).

The RoB 2 assesses five key domains that can introduce bias in RCTs:

i. Bias arising from the randomization process.
ii. Bias due to deviations from intended interventions.
iii. Bias due to missing outcome data.
iv. Bias in the measurement of the outcome.
v. Bias in the selection of the reported result.

The risk of bias will be categorized into three levels. ‘Low Risk’ indicates that the trial is unlikely to be influenced by bias in the assessed domain. ‘Some Concerns’ suggests that there is a possibility of bias that could reduce confidence in the results. ‘High Risk’ implies that bias in the domain is likely to significantly impact the reliability of the findings.

### 5.4. Data Synthesis

The systematic review will provide a comprehensive overview of the included studies. Data extraction will focus on self-help interventions, the outcomes measured in relation to these interventions, and their corresponding results. This process involved synthesizing the findings of the available studies and assessing the certainty and significance of the identified outcomes. Findings will be summarized, with results presented in tabular format. The quality of evidence will be evaluated based on the GRADE framework, considering factors such as study design, risk of bias, precision, inconsistency, indirectness, and effect size. Evidence certainty will be categorized into four levels: “High,” “Moderate,” “Low,” and “Very Low” (21).

## 6. Discussion

Complex interventions for managing mild to moderate depression are characterized by multiple interacting components, contextual dependencies and varied outcomes (22, 23). Theory-based interventions often integrate elements such as psychoeducation, cognitive restructuring and behavioral activation, with features to enhance user engagement (24). Evaluating the individual and combined effects of these components, along with understanding user interactions with the digital platform and contextual influences like cultural norms and healthcare infrastructure, is crucial for assessing their effectiveness, adaptability, and scalability (25, 26). This systematic review will address these dimensions of complexity, the component analysis, interaction assessment and contextual evaluation, to provide a nuanced understanding of the factors influencing the success of theory-based interventions for depression. The findings will offer actionable insights for optimizing complex interventions’ design, implementation and scalability.

### Health Equity Considerations

Incorporating health equity considerations is essential for evaluating digital interventions for mild to moderate depression. Health equity refers to eliminating avoidable disparities in health outcomes across population groups, influenced by factors like socioeconomic status, geographic location, ethnicity and access to resources (27). Digital interventions have the potential to overcome barriers such as geographic isolation and limited healthcare access, expanding their reach to underserved populations (28). However, disparities in digital literacy, internet access and the cultural relevance of intervention content may hinder equitable distribution of benefits (29, 30). This review will assess the extent to which included studies address health equity by examining population characteristics (place of residence, ethnicity, occupation, gender, religion, education, social capital and socio-economic position) (31). Additionally, given the variability in individual responses, not all theoretical frameworks are equally effective for all users, necessitating careful adaptation and customization of interventions (32).

### Limitations and Added Value

A notable limitation of this review may be measurement bias from the common use of self-reporting of outcomes or self-declared as having depression, without undergoing clinical assessment or receiving a confirmed diagnosis from a psychiatrist or other qualified mental health professional. This reliance may limit the certainty and comparability of findings (32). Despite this limitation, the review offers significant contributions to the field of mental health interventions. It provides a detailed analysis of interventions rooted in psychological theories, elucidating the foundational principles that underpin their design and effectiveness (34). By exploring delivery methods such as web-based platforms and mobile applications, the review offers a detailed insights of how delivery formats influence engagement, adherence and outcomes, catering to diverse user preferences (35, 36).

## Conclusion

In conclusion, this systematic review will provide a comprehensive evaluation of the role of self-help practices derived from theory-based intervention in managing adults with mild to moderate depression. The findings will support the integration of theory-based intervention as a viable non-pharmacological option, offering long-term benefits such as symptom reduction, relapse prevention and improved quality of life. Our review will highlight the novel findings where there may be certainty of theory-based interventions to reduce depressive symptoms severity. These results have profound implications for guiding global health practice and policy to mental health interventions in primary care.

## Data Availability

N/A

## Author Contributions

**Conceptualization:** Asma’ Khalil, Chen Xin Wee.

**Methodology:** Asma’ Khalil, Chen Xin Wee, Philip Baker.

**Project Administration:** Asma’ Khalil, Chen Xin Wee

**Supervision and Validation:** Chen Xin Wee, Zahir Izuan Azhar, Norley Shuib, Philip Baker.

**Writing– Original Draft Preparation:** Asma’ Khalil.

**Writing– Review & Editing:** Asma’ Khalil, Chen Xin Wee, Zahir Izuan Azhar, Philip Baker.

